# Epilepsy Surgery vs Medical Management for Pediatric Drug-Resistant Focal Epilepsy

**DOI:** 10.64898/2026.07.10.26357665

**Authors:** Taylor J. Abel, Emily Harford, Daniel Silliman, Ruba Al-Ramadhani, Samuel Wiebe, Kenneth J. Smith

## Abstract

**Background and Objectives:** Drug-resistant focal epilepsy affects approximately 30% of children with epilepsy and carries excess mortality, impaired neurodevelopment, and substantial costs. Epilepsy surgery is underutilized despite proven superiority over medical management and MRI-guided laser interstitial thermal therapy (MRgLITT) is a minimally invasive alternative to open resection, but comparative evidence to guide procedure selection is limited. Our objective was to estimate lifetime outcomes and costs of epilepsy surgery versus medical management for pediatric drug-resistant focal epilepsy, and to provide etiology-informed guidance for choosing between open resection and MRgLITT.

**Methods:** We developed a Markov-based decision analytic model over a lifetime horizon to compare best medical management, open resection, and MRgLITT for drug-resistant focal epilepsy from a US healthcare payer perspective. The base case was a 10-year-old child with a seizure focus of <3 cm³. Model input parameters were obtained from published systematic reviews and meta-analyses. The primary outcome was the incremental cost-effectiveness ratio, expressed as cost per quality-adjusted life years gained. Probabilistic and deterministic sensitivity analyses were conducted to evaluate uncertainty.

**Results:** Both surgical strategies were associated with a 4.6-year survival advantage, 3.6 additional lifetime QALYs, and lower costs than medical management. MRgLITT yielded 22.64 QALYs at $120,943; open resection yielded 22.62 QALYs at $121,649; medical management yielded 19.00 QALYs at $127,465. The difference between MRgLITT and open resection was 0.015 QALYs, reflecting near-equivalent effectiveness; in probabilistic sensitivity analysis, MRgLITT was optimal in 50.3% of iterations and open resection in 38.3%, with neither showing clear superiority. Etiology-specific analyses favored MRgLITT for focal cortical dysplasia and mesial temporal sclerosis, and open resection for tumor-related and cavernoma-related epilepsy.

**Discussion:** Both open resection and MRgLITT were associated with substantially better lifetime outcomes and lower costs than medical management, supporting early surgical referral. Overall effectiveness between surgical approaches was clinically similar, with neither demonstrating clear superiority; the model suggests epilepsy etiology, rather than expected effectiveness alone, should guide procedure selection between MRgLITT and open resection.

## Introduction

Drug-resistant epilepsy (DRE), defined as failure of two appropriate antiseizure medications to achieve seizure freedom^1^, affects approximately 30% of children with epilepsy. For children with a focal seizure onset, epilepsy surgery offers the opportunity for seizure freedom and its benefits, which include improved neurodevelopmental trajectory^2^, reduced epilepsy-related mortality^3,4^, and enhanced quality of life^5–8^. Despite level I evidence supporting surgical superiority over continued medical management in both adults and children^9,10^, surgical intervention remains substantially underutilized in children^11–13^, with estimates suggesting that only a small proportion of candidates annually undergo surgery. Delays in referral lead to continued seizures, persistent neurodevelopmental impairment, increased mortality risk, and cumulative costs^14^.

Open resective surgery has historically been the standard surgical approach for drug-resistant focal epilepsy, achieving seizure freedom in approximately 60–65% of pediatric patients^15–20^. MRI-guided laser interstitial thermal therapy (MRgLITT) has emerged as a minimally invasive alternative and is now widely used at pediatric epilepsy surgery centers, offering advantages including smaller incision, shorter hospitalization, and suitability for deep or eloquent cortex targets^21–23^. Reported seizure freedom rates range from approximately 48–65% depending on etiology and lesion characteristics^15,16,23–25^, which is comparable to open resection in some contexts, but potentially inferior in others. Recently, a retrospective cohort analysis by Yossofzai et al.^17^ showed inferiority of MRgLITT for seizure control, but this analysis does not holistically compare surgical outcomes beyond seizure control measures. Despite the rapid adoption of MRgLITT in clinical practice, comparative evidence to guide procedure selection in children is limited, and no head-to-head trials exist. Clinicians and families currently choose between these interventions without a systematic framework for weighing the tradeoffs across the lifetime horizon, which matters most in pediatric disease.

Economic analyses of epilepsy surgery in adults have demonstrated favorable long-term outcomes and costs^26^, but the pediatric context is unique due to longer time horizons that amplify both benefits and the consequences of seizure control and related costs. A randomized trial comparing open resection to MRgLITT in children broadly across focal epilepsies is unlikely to be feasible given sample size requirements, the ethical challenges of surgical randomization, and the rapid pace of adoption that has already established both approaches in practice.

We therefore developed a decision analytic model to address two clinical questions: first, what is the lifetime consequence, in survival, quality-adjusted life, and projected cost, of epilepsy surgery versus continued best medical management for children with drug-resistant focal epilepsy? Second, when both open resection and MRgLITT are technically feasible, how should clinicians choose between them? We focused on patients with a seizure focus of <3 cm³ - per Yossofzai et al.^17^ - to ensure clinical comparability between approaches and performed etiology-specific scenario analyses for common focal epilepsy etiologies in children.

## Methods

We developed a Markov-based decision analytic model to compare three treatment strategies for pediatric drug-resistant focal epilepsy: 1) best medical management, 2) open seizure focus resection, and 3) MRgLITT. Based on a recent comparative effectiveness study^17^, the base case was a 10-year-old child with drug-resistant focal epilepsy and a seizure focus of <3 cm³. A maximal seizure focus of <3 cm³ was chosen to ensure clinical equipoise between open resection and MRgLITT, given the feasibility of ablating larger volumes with the latter. Drug-resistant epilepsy was defined per the International League Against Epilepsy as failure of two appropriate antiseizure medications to achieve seizure freedom^1^. The analysis adopted a healthcare payer perspective in the United States over a lifetime time horizon.

Model outcomes included quality-adjusted life years (QALYs), direct medical costs (2025 US dollars), lifetime survival, and incremental cost-effectiveness ratios (ICERs). All costs were inflated to 2025 US dollars using the Personal Consumption Expenditure Index. Future costs and QALYs were discounted at 3% annually per Office of Management and Budget guidelines^27^. This study followed the Consolidated Health Economic Evaluation Reporting Standards (CHEERS) reporting guidelines.

### Model Development

A schematic of the decision model is presented in Figure 1. Models were built and analyzed using TreeAge Pro software (TreeAge Software, Plymouth, MA). Each strategy was represented as a Markov model with a 1-year cycle length and a lifetime time horizon beginning at age 10 years. The model tracked patients through health states defined by seizure control status^28^ (seizure free [ILAE class 1–2] or disabling seizures [ILAE class 3–6]) and the presence or absence of a permanent neurologic deficit. An absorbing dead state was included in all models. For surgical strategies (open versus MRgLITT), initial model branches captured the probability of perioperative mortality, major surgical complication, and initial seizure outcome. Patients not achieving seizure freedom after initial surgery could undergo revision surgery at standard rates observed in the literature^29^, modeled as open resection regardless of the index procedure. Surgical complications without permanent neurologic deficit were modeled via a transient disutility and cost applied in the cycle of occurrence. Annual cycle transitions captured long-term changes in seizure control, including seizure recurrence among patients who achieved freedom and seizure remission among those who did not. The model was run until cohort exhaustion; the median age at death of 60–64 years in the base case confirms that results do not depend on extrapolation to implausible survival ages. Health state–specific utilities and standardized mortality ratios were applied each cycle to accumulate QALYs and estimate survival, respectively. Age-specific background mortality was derived from 2021 US National Center for Health Statistics life tables^30^ and adjusted using epilepsy-specific standardized mortality ratios.

**Figure 1.**
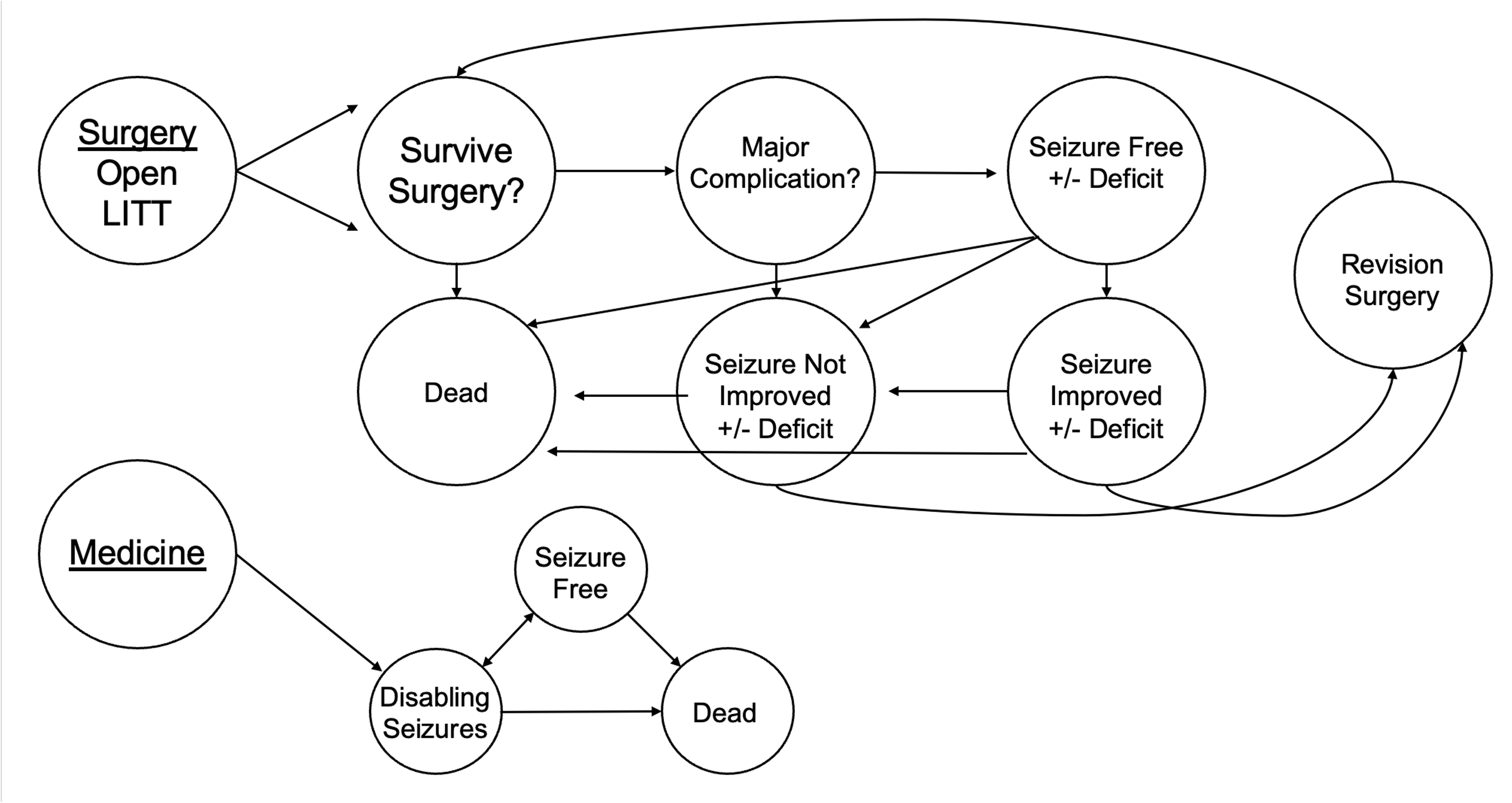
Markov Decision Analytic Model Structure. Schematic of the Markov-based decision analytic model comparing open resection, MRI-guided laser interstitial thermal therapy (MRgLITT), and best medical management for pediatric drug-resistant focal epilepsy. Surgical strategies share a common decision tree capturing perioperative mortality, major complications, initial seizure outcome, and revision surgery, followed by annual Markov cycle transitions between seizure health states. The medical management arm models annual transitions between seizure-free and disabling seizure states. Health state–specific utilities and standardized mortality ratios are applied each annual cycle.

### Input Parameters

Model input parameters, probabilistic sensitivity analysis ranges, and distributions are summarized in Table 1 and sources used to derive parameters are listed in Supplementary Table 1. Clinical probabilities for surgical outcomes, including complication rates, permanent neurologic deficit rates, seizure freedom rates, and revision surgery rates, were derived from published systematic reviews and meta-analyses with preference for multi-center studies and larger sample sizes. For surgical outcome parameters (complication rates, seizure freedom rates, and revision surgery rates), base case values represent weighted pooled estimates across multiple sources and the range across sources informed probabilistic sensitivity analysis distributions. Transition probabilities and health state utilities were each derived from a single best-available source^31,32^, and standardized mortality ratios were derived from published long-term epilepsy outcome studies (Supplementary Table 1). Seizure recurrence and remission transition probabilities were obtained from a longitudinal cohort study of postoperative seizure outcomes^31^. The probability of seizure freedom with best medical management was derived as the complement of the weighted probability of continued disabling seizures reported in available literature. Etiology-specific parameters for scenario analyses were obtained from published cohort studies and meta-analyses and are also presented in Supplementary Table 1.

**Table 1.**
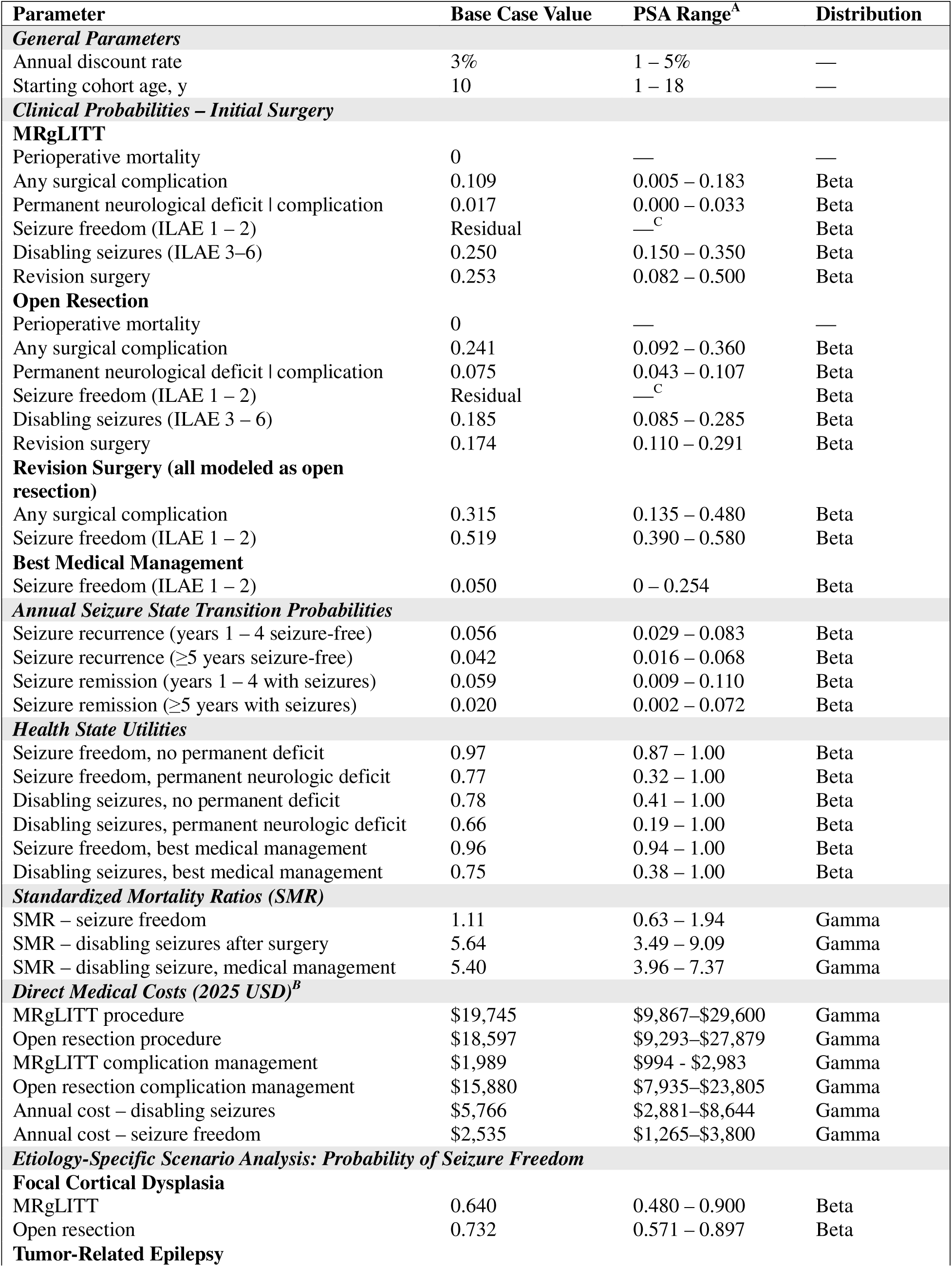
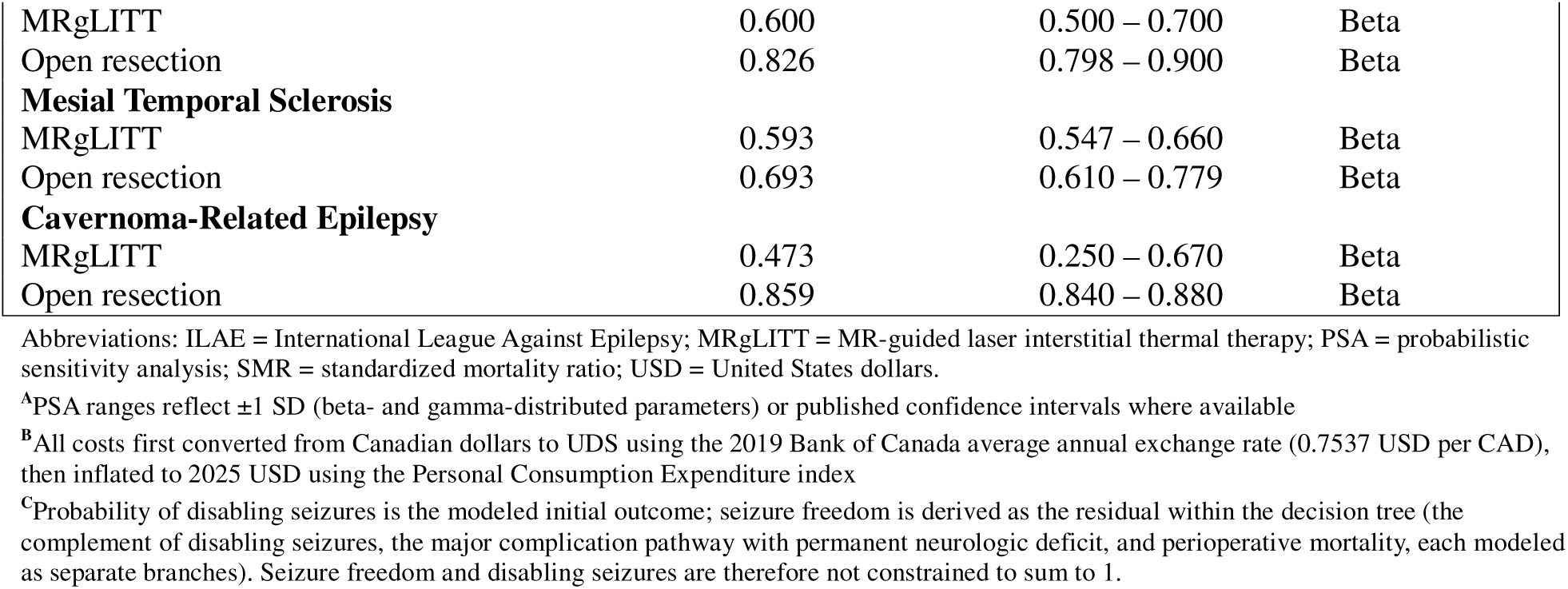
Model Input Parameters for Markov Decision Analysis of Pediatric Focal Drug-Resistant Epilepsy.

Direct medical costs included procedure costs, complication management costs, and annual health state– specific costs of seizure care and antiseizure medication. Procedure and complication costs were obtained from published cost analyses and adjusted to 2025 US dollars using the Personal Consumption Expenditure Index. Because the source cost data were reported in Canadian dollars, all costs were first converted to US dollars using the 2019 Bank of Canada average annual exchange rate (0.7537 USD per CAD) and then inflated to 2025 US dollars using the Personal Consumption Expenditure Index (inflation factor 1.2222). Annual costs of ongoing seizure-related care were similarly inflated to 2025 US dollars.

### Analysis

Strategies were ranked by total cost and effectiveness, and incremental cost-effectiveness ratios were calculated for non-dominated strategies using standard methods. Strategies subject to strong or extended dominance were excluded from the ICER calculation. A willingness-to-pay threshold of $100,000 per QALY was used as the primary reference in accordance with commonly applied US standards, with secondary reference to $150,000 and $200,000 per QALY thresholds.

One-way deterministic sensitivity analyses were performed for all model parameters over their plausible ranges. Results are presented as incremental net monetary benefit (NMB) tornado diagrams comparing each strategy pair, with NMB calculated at a $100,000/QALY willingness-to-pay threshold. NMB was used in preference to ICER to avoid undefined values when dominance relationships are present. Parameters were ranked by the absolute range in incremental NMB across their sensitivity analysis bounds. A deterministic time horizon sensitivity analysis was also performed to test whether the base case findings depended on the lifetime horizon.

Probabilistic sensitivity analysis (PSA) was performed using Monte Carlo simulation with 10,000 iterations. Clinical probabilities were assigned beta distributions; costs were assigned gamma distributions; and standardized mortality ratios were assigned gamma distributions. Distribution parameters were derived from published confidence intervals or calculated from base case means and assumed standard deviations as detailed in Table 1. Results are presented as incremental cost-effectiveness scatterplots and cost-effectiveness acceptability curves (CEACs) showing the probability that each strategy was optimal across willingness-to-pay thresholds from $0 to $200,000 per QALY.

Markov cohort survival analysis was performed to estimate the survival benefit of each strategy. Mean survival and median age at death were calculated from the cohort survival curves for each strategy.

We performed etiology-specific scenario analyses to evaluate cost-effectiveness across epilepsy subtypes most commonly treated surgically: focal cortical dysplasia (FCD), tumor-related epilepsy, cavernoma-related epilepsy, and mesial temporal sclerosis (MTS). For each etiology, published seizure freedom rates specific to that etiology were substituted for the base case values, with all other parameters held at base case values. Disabling seizure probabilities were calculated as the complement of the etiology-specific seizure freedom rate and the base case revision surgery rate.

## Results

### Base Case Analysis

In the base case analysis, both surgical strategies were associated with a 4.6-year mean survival advantage (51.6 years for surgery vs. 47.0 years for medical management; median age at death 64 vs. 60 years), approximately 3.6 additional lifetime QALYs, and lower projected lifetime costs. MRgLITT yielded 22.64 QALYs at a projected lifetime cost of $120,943; open resection yielded 22.62 QALYs at $121,649; and best medical management yielded 19.00 QALYs at $127,465. The difference in effectiveness between MRgLITT and open resection was 0.015 QALYs, which is equivalent to approximately 5 quality-adjusted days over a lifetime, reflecting the near-equivalent long-term effectiveness. Complete base case results are presented in Table 2.

**Table 2.**
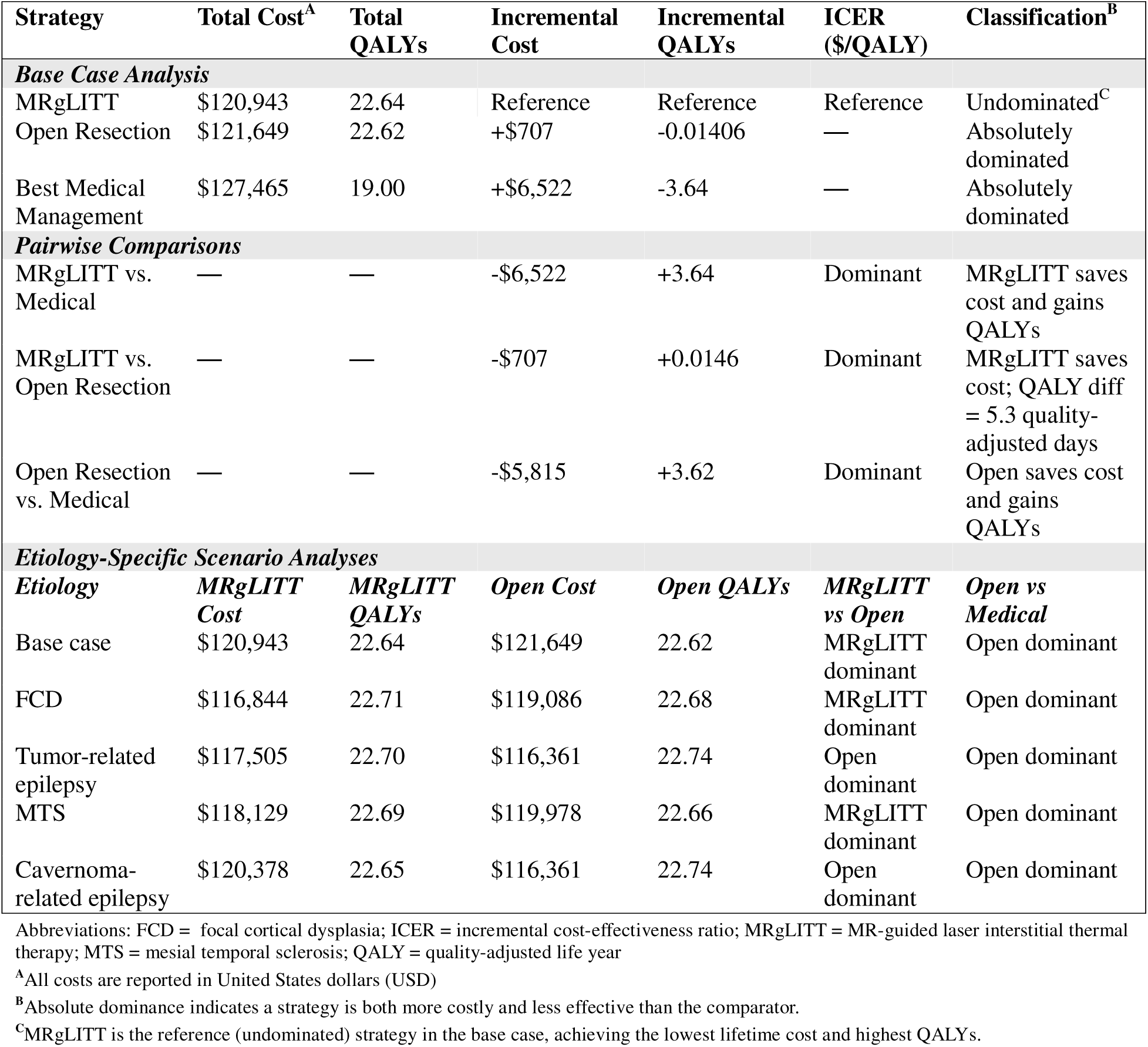
Base Case Cost-Effectiveness Analysis of Treatment Strategies for Pediatric Focal Drug-Resistant Epilepsy.

The survival benefit of surgery reflects lower epilepsy-specific standardized mortality ratios in the seizure-free state, compounded over a lifetime beginning at age 10. Projected lifetime healthcare costs were lower for both surgical strategies than for best medical management (by $5,815–$6,522), because the upfront procedural investment is offset by sustained reductions in seizure-related healthcare utilization when seizure freedom is achieved early in childhood.

### Deterministic Sensitivity Analysis

Results of one-way deterministic sensitivity analyses are presented in Figure 2. For the comparison of open resection versus best medical management (Figure 2A), the probability of seizure freedom with medical management was the most influential parameter, followed by the discount rate and probability of seizure recurrence after ≥5 years of seizure freedom. Procedure costs and complication rates had comparatively little influence on this comparison, indicating that the result is driven primarily by the effectiveness difference rather than cost differences between strategies.

**Figure 2.**
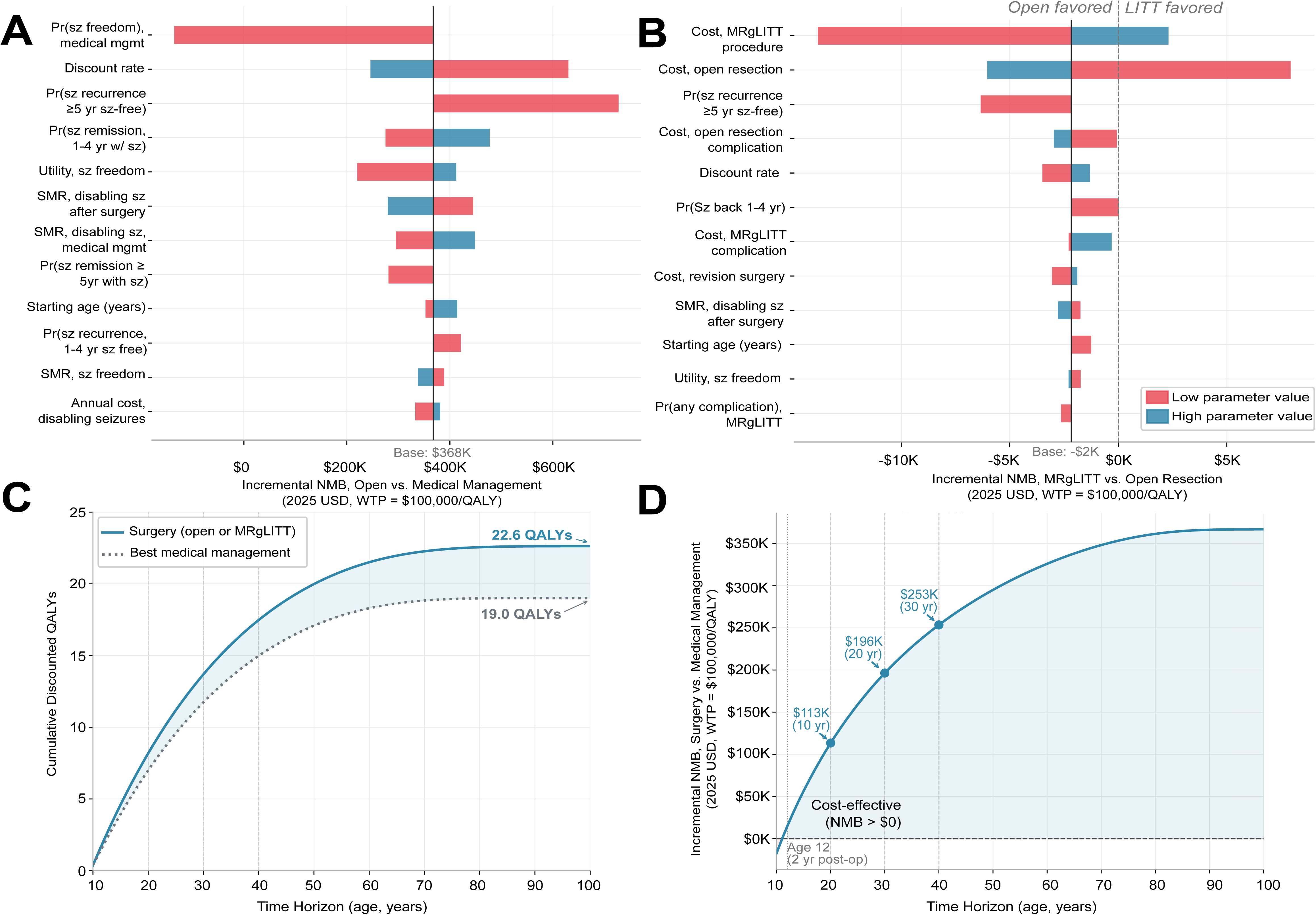
One-Way Deterministic Sensitivity Analysis. Tornado diagrams depicting the influence of individual parameter variation on incremental net monetary benefit (NMB) at a willingness-to-pay threshold of $100,000 per quality-adjusted life year (QALY). Parameters are ranked by the absolute range in incremental NMB across their plausible bounds, with the most influential parameter at the top. The vertical line represents the base case incremental NMB. Blue bars indicate the NMB when the parameter is at its low value; red bars indicate the NMB when the parameter is at its high value. **Panel A** shows the comparison of open resection versus best medical management (base case incremental NMB = $367K). **Panel B** shows the comparison of MRgLITT versus open resection (base case incremental NMB = −$2K; negative value indicates MRgLITT is favored). The dashed vertical line in Panel B at zero NMB represents the threshold at which the preferred strategy switches. **Panel C** compares discounted QALYs for surgery versus best medical management in a 100-year time horizon, with a 3.6 cumulative discounted QALY advantage for surgery. **Panel D** compares cost effectiveness of surgery versus best medical management on the same time horizon, with a projected improved cost effectiveness for surgery within 2 years post-op.

For the comparison of MRgLITT versus open resection (Figure 2B), procedure costs were the dominant parameters: the costs of the MRgLITT and open resection procedures were the two most influential variables. Several parameters including procedure costs and starting age crossed the zero incremental NMB threshold, indicating that under certain parameter combinations the preferred strategy switches between MRgLITT and open resection. This threshold-crossing behavior is consistent with the near-equivalent base case effectiveness of the two strategies: because effectiveness differences are small, cost differences become the primary driver of relative value, and the outcome is sensitive to the relative pricing of each procedure. Although revision surgery rates differed between MRgLITT (25.3%) and open resection (17.4%), this parameter was not an influential driver of model outcomes: the cost of revision surgery ranked 17th of 19 parameters for the open resection versus best medical management comparison (0.5% of maximum parameter influence) and 8th of 21 parameters for the MRgLITT versus open resection comparison, reflecting that modeled outcomes were driven primarily by procedure costs, initial seizure freedom rates, long-term seizure recurrence probabilities, and mortality.

Because long-term outcome data for pediatric focal epilepsy surgery remain sparse, we conducted a deterministic time horizon sensitivity analysis to test whether the base case findings depended on the lifetime horizon used in the primary analysis. Extending the model to a lifetime time horizon and discounting future QALYs at the standard 3% annual rate, surgery (MRgLITT or open resection) retained a 3.6 cumulative discounted QALY advantage over best medical management, closely matching the base case estimate and confirming that the QALY benefit is not an artifact of the finite time horizon used in the primary analysis (Figure 2C). Surgery also became cost-effective relative to best medical management within 2 years of the index procedure across this extended horizon, indicating that the economic advantage of surgery emerges early after surgery and is not contingent on model parameters for extended follow-up (Figure 2D).

### Probabilistic Sensitivity Analysis

Results of the probabilistic sensitivity analysis are presented in Figure 3. The incremental cost-effectiveness scatterplot for surgery versus best medical management (Figure 3A) confirmed robust surgical benefit: open resection was cost-effective at a $100,000/QALY willingness-to-pay threshold in 87.0% of iterations, with the 95% confidence ellipse situated predominantly in the favorable quadrant indicating lower cost and greater effectiveness. The scatterplot for MRgLITT versus open resection (Figure 3B) told a different story: the distribution was tightly clustered near the origin of the cost-effectiveness plane, reflecting the near-equivalence of the two surgical strategies. MRgLITT was marginally favored (cost-effective) in 46.0% of iterations at the $100,000/QALY threshold — statistically indistinguishable from open resection and consistent with genuine clinical equipoise between approaches.

**Figure 3.**
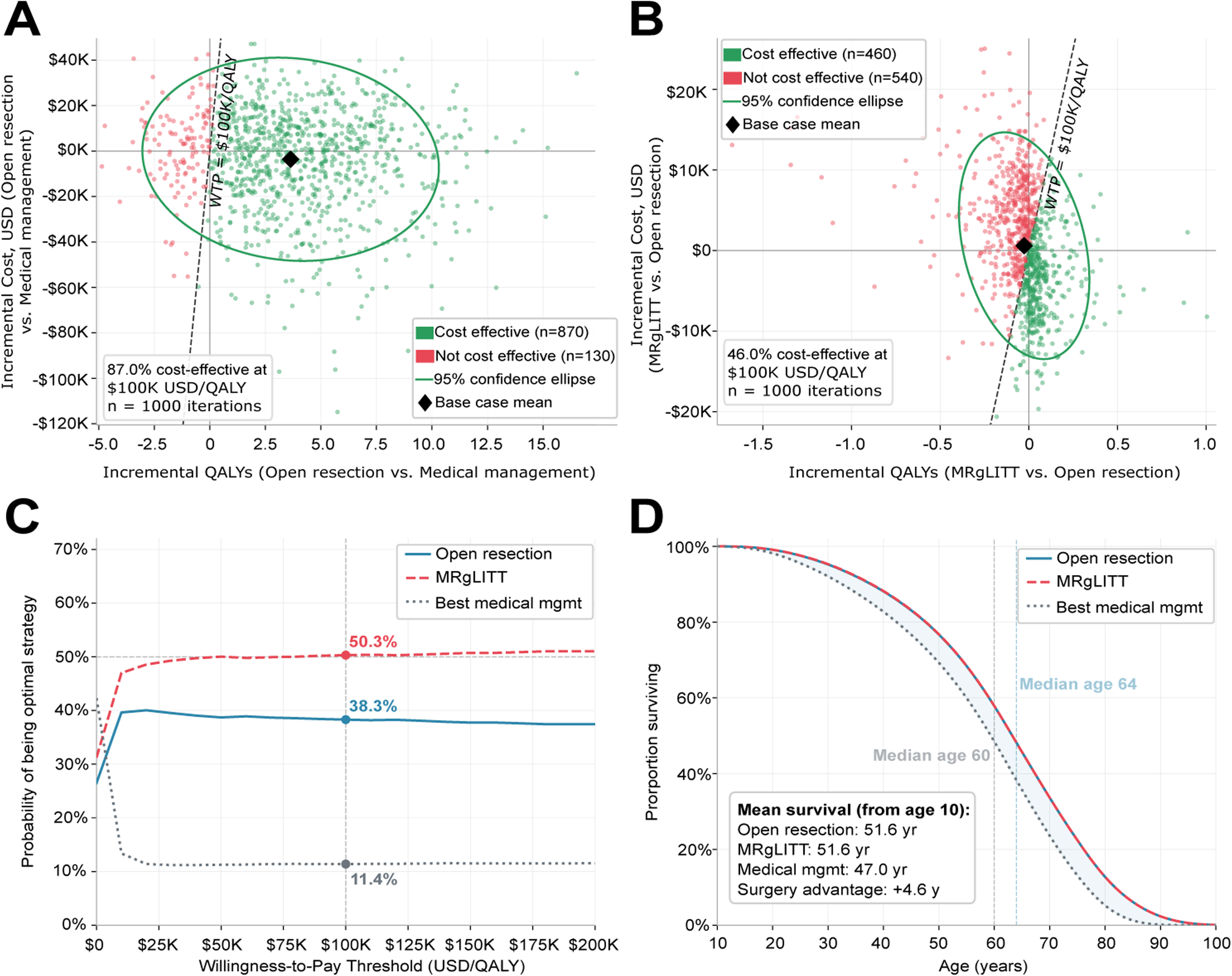
Probabilistic Sensitivity Analysis and Survival. Results of Monte Carlo probabilistic sensitivity analysis (10,000 iterations) and Markov cohort survival analysis. **Panel A:** Incremental cost-effectiveness scatterplot for open resection versus best medical management. Each point represents one PSA iteration. Green points are cost-effective at a $100,000/QALY willingness-to-pay threshold; red points are not cost-effective. The ellipse denotes the 95% confidence region; the diamond denotes the base case mean. Open resection was cost-effective in 87.0% of iterations. **Panel B:** Incremental cost-effectiveness scatterplot for MRgLITT versus open resection. The tightly clustered distribution centered near the origin reflects near-equivalent effectiveness between strategies. MRgLITT was cost-effective in 46.0% of iterations. **Panel C:** Cost-effectiveness acceptability curves showing the probability that each strategy is optimal across willingness-to-pay thresholds from $0 to $200,000/QALY. At $100,000/QALY, MRgLITT was optimal in 50.3% of iterations, open resection in 38.3%, and best medical management in 11.4%. **Panel D:** Markov cohort survival curves for each strategy. The shaded region represents the survival advantage of surgery over best medical management.

Cost-effectiveness acceptability curves (Figure 3C) demonstrated that at a $100,000/QALY willingness-to-pay threshold, MRgLITT was the optimal strategy in 50.3% of iterations, open resection in 38.3%, and best medical management in only 11.4%. The near-equal probabilities of optimality for MRgLITT and open resection across willingness-to-pay thresholds reinforce that neither surgical approach has clear overall superiority under parameter uncertainty. Importantly, best medical management was the optimal strategy in fewer than 15% of iterations above a $10,000/QALY threshold across the full range examined, confirming that the clinical benefit of epilepsy surgery over continued medical management is robust to joint parameter uncertainty.

### Survival Analysis

Markov cohort survival analysis demonstrated a meaningful survival advantage for surgical treatment over best medical management (Figure 3D). Mean survival from age 10 was 51.6 years for both surgical strategies, compared with 47.0 years for best medical management, representing a 4.6-year surgery-associated advantage. The survival curves for open resection and MRgLITT were nearly overlapping throughout the model horizon, reflecting their near-equivalent seizure freedom rates (Table 1). The survival benefit of surgery is attributable to lower standardized mortality ratios in the seizure-free health state, compounded over the lifetime.

### Etiology-Specific Scenario Analyses

Etiology-specific scenario analyses demonstrated that both surgical strategies were less costly and more effective than best medical management across all epilepsy subtypes examined (Table 2). The preferred surgical strategy, however, varied by etiology according to the magnitude of the seizure freedom difference between MRgLITT and open resection. In focal cortical dysplasia and mesial temporal sclerosis etiologies, in which the difference in seizure freedom rates between MRgLITT and open resection is relatively small (0.083 and 0.100, respectively), MRgLITT was marginally favored in the base case model. In FCD, MRgLITT yielded 22.71 QALYs at $116,844 compared with 22.68 QALYs at $119,086 for open resection. In MTS, MRgLITT yielded 22.69 QALYs at $118,129 compared with 22.66 QALYs at $119,978 for open resection. In both etiologies, the modestly lower projected procedural cost of MRgLITT outweighed its marginally lower seizure freedom rate, though the absolute QALY differences between approaches were small (0.03 QALYs or less). In technically appropriate cases where the lesion is amenable to ablation, MRgLITT may represent a reasonable first approach in these etiologies.

In tumor-related epilepsy and cavernoma-related epilepsy, etiologies with larger absolute differences in seizure freedom rates between open resection and MRgLITT (0.226 and 0.386, respectively), open resection was the preferred strategy in the model. In tumor-related epilepsy, open resection yielded 22.74 QALYs at $116,361 compared with 22.70 QALYs at $117,505 for MRgLITT. In cavernoma-related epilepsy, the difference was most pronounced: open resection yielded 22.74 QALYs at $116,361 compared with 22.65 QALYs at $120,378 for MRgLITT, reflecting the large gap in modeled seizure freedom rates in this etiology (0.859 for open resection vs. 0.473 for MRgLITT). In these etiologies, the substantially higher seizure freedom associated with open resection outweighs any procedural cost advantage of MRgLITT, and the model supports open resection as the preferred approach when technically feasible.

## Discussion

In this decision analytic model of pediatric drug-resistant focal epilepsy, surgery was associated with a modeled 4.6-year survival advantage, approximately 3.6 additional lifetime QALYs, and lower projected lifetime healthcare costs compared with best medical management. These findings were consistent across all base case and etiology-specific scenarios. The projected lifetime cost advantage of surgery ($5,815–$6,522 compared with medical management) reflects the sustained reduction in seizure-related healthcare utilization when seizure freedom is achieved in childhood, which offsets the upfront procedural costs over a lifetime horizon. Prior analyses in adults have similarly demonstrated favorable economics of surgery^26^, but the pediatric context is crucial to consider given the long-term neurodevelopmental and cost consequences of improved seizure control with surgery. These findings provide quantitative support for early surgical referral and underscore the underutilization of epilepsy surgery in children.

The near-equivalent overall effectiveness of MRgLITT and open resection, which differed by only 0.015 QALYs over a lifetime, is a central finding of our analysis. Because overall lifetime effectiveness is similar, the choice between approaches should be driven primarily by epilepsy etiology, lesion characteristics, and patient and family preferences rather than expected effectiveness alone. The etiology-specific analyses demonstrate that this near-equivalence may not be uniform. For example, our findings suggest that MRgLITT may be a reasonable first approach in focal cortical dysplasia and mesial temporal sclerosis, where modeled seizure freedom rates are relatively similar between approaches. Meanwhile, open resection may be preferred in tumor-related and cavernoma-related epilepsy, where it is associated with substantially higher modeled seizure freedom. This pattern is consistent with emerging evidence that MRgLITT outcomes vary substantially by etiology and lesion type^17,37^. These findings should be interpreted as a model-based decision aid and not a clinical dogma as individual anatomy, surgeon experience or preference, and patient preference should be considered in surgical decision making.

The finding that MRgLITT is the modeled dominant strategy in the base case and in FCD and MTS appears counterintuitive given prior evidence suggesting generally inferior seizure outcomes with MRgLITT^15,16,38^. However, the dominance of MRgLITT does not arise from superior seizure control; in the base case, open resection achieves meaningfully higher effective seizure freedom (a modeled 64.1% vs 49.7%, derived from disabling-seizure probabilities of 0.185 vs 0.250; Table 1). Rather, it reflects MRgLITT’s substantially lower complication rate (10.9% vs 24.1%) and dramatically lower complication management cost ($1,989 vs $15,880), which together reduce expected complication costs by approximately $3,600. Combined with MRgLITT’s lower permanent neurologic deficit rate (1.7% vs 7.5%), which avoids a persistent health state utility penalty over a lifetime beginning at age 10, these factors tip the long-term balance toward MRgLITT in etiologies where the seizure freedom gap is relatively small. In tumor-related and cavernoma-related epilepsy, where open resection achieves substantially higher seizure freedom (differences of 0.226 and 0.386, respectively), the effectiveness advantage overwhelms MRgLITT’s cost and complication advantages. The findings highlight that the choice between approaches is not simply about seizure freedom rates, but also needs to incorporate complication profiles, permanent neurologic sequelae, and their interaction with lifetime health state utilities. The probabilistic sensitivity analysis reinforces that neither surgical approach has clear overall superiority under joint parameter uncertainty. MRgLITT was the optimal strategy in 50.3% of PSA iterations and open resection in 38.3% at a $100,000/QALY threshold, which is a narrow difference that reflects equipoise rather than meaningful superiority for either approach. This uncertainty accurately reflects the current evidence base: long-term pediatric MRgLITT data are still accumulating, and the relative balance between approaches may shift as larger comparative series mature. These limitations reinforce the appropriateness of individualized, etiology-informed decision-making rather than application of a single universal protocol. That said, if emerging evidence shows specifically neurodevelopmental or quality of life benefit for one approach or another, as suggested by prior work on MRgLITT in language-dominant MTLE^39,40^, one strategy may become favored in specific contexts.

Several limitations warrant consideration. First, the model was parameterized primarily from observational studies and meta-analyses rather than randomized trial data, and heterogeneity and potential selection bias in surgical series introduce uncertainty captured only partially in sensitivity analyses. Second, long-term pediatric MRgLITT outcome data are limited; reported series are generally smaller and have shorter follow-up than open resection series, which may introduce imprecision in MRgLITT seizure freedom estimates. Third, antiseizure medication side effects were not modeled as a separate health state; however, utility variation across probabilistic sensitivity analysis ranges encompasses the magnitude of utility decrement associated with medication side effects reported in published epilepsy cohorts. To the extent that best medical management is associated with higher ongoing medication burden, omitting explicit side effect modeling may modestly underestimate the relative benefit of surgery. Fourth, the model does not capture neurodevelopmental outcomes, cognitive gains, or quality-of-life improvements beyond those reflected in health state utilities, which likely underestimates the true benefits of early seizure freedom in children, particularly for educational attainment and long-term independence. Fifth, indirect costs were not incorporated, consistent with the healthcare payer perspective adopted for this analysis. Sixth, although our time horizon sensitivity analyses projected improved QALYs and cost effectiveness for surgical interventions over relatively short time horizons (Fig. 2C/D), longer term results are modeled based on wide assumptions regarding parameters. Finally, costs and utilities were derived from published sources and may not reflect contemporary practice patterns or institutional variation in procedural costs and complication rates. Additionally, health state utilities and standardized mortality ratios were derived from adult epilepsy cohorts and may not fully reflect the preferences and mortality experience of children with epilepsy, for whom neurodevelopmental consequences of seizures and health state values may differ substantially from adults.

## Conclusion

In this decision analytic model, both open resection and MRgLITT were associated with substantially better modeled lifetime outcomes and lower projected costs than best medical management for pediatric drug-resistant focal epilepsy, supporting early surgical referral for appropriate candidates. Between surgical approaches, overall lifetime effectiveness was clinically similar; the model suggests that epilepsy etiology should guide procedure selection, with MRgLITT as a reasonable consideration in focal cortical dysplasia and mesial temporal sclerosis, and open resection preferred in tumor-related and cavernoma-related epilepsy. These findings provide a quantitative, etiology-informed framework to support individualized shared decision-making, and reinforce the urgency of timely surgical evaluation for children with drug-resistant focal epilepsy.

## Data Sharing Statement

No new patient-level data were generated for this study. All model input parameters were derived from previously published sources, cited in Table 1. The TreeAge Pro decision-analytic model file is available from the corresponding author upon reasonable request.

## Data Availability

All data produced in the present work are contained in the manuscript. No new patient-level data were generated as a result of the present study.

## References

1. Kwan, P. et al. Definition of drug resistant epilepsy: Consensus proposal by the ad hoc Task Force of the ILAE Commission on Therapeutic Strategies. Epilepsia 51, 1069–1077 (2010).

2. Stefanos-Yakoub, I. et al. Long-term intellectual and developmental outcomes after pediatric epilepsy surgery: A systematic review and meta-analysis. Epilepsia 65, 251–265 (2024).

3. Granthon, C. et al. Reduced long-term mortality after successful resective epilepsy surgery: a population-based study. J Neurol Neurosurg Psychiatry 95, 249–255 (2024).

4. Bell, G. S. et al. Premature mortality in refractory partial epilepsy: does surgical treatment make a difference? *Journal of Neurology*, Neurosurgery & Psychiatry 81, 716–718 (2010).

5. Widjaja, E. et al. Trajectory of Health-Related Quality of Life After Pediatric Epilepsy Surgery. JAMA Netw Open 6, e234858 (2023).

6. Mikati, M. A. et al. Quality of life after surgery for intractable partial epilepsy in children: a cohort study with controls. Epilepsy Res 90, 207–213 (2010).

7. Van Empelen, R., Jennekens-Schinkel, A., Van Rijen, P. C., Helders, P. J. M. & Van Nieuwenhuizen, O. Health-related Quality of Life and Self-perceived Competence of Children Assessed before and up to Two Years after Epilepsy Surgery. Epilepsia 46, 258–271 (2005).

8. Fiest, K. M., Sajobi, T. T. & Wiebe, S. Epilepsy surgery and meaningful improvements in quality of life: Results from a randomized controlled trial. Epilepsia 55, 886–892 (2014).

9. Wiebe, S., Blume, W. T., Girvin, J. P. & Eliasziw, M. A Randomized, Controlled Trial of Surgery for Temporal-Lobe Epilepsy. New England Journal of Medicine 345, 311–318 (2001).

10. Dwivedi, R. et al. Surgery for Drug-Resistant Epilepsy in Children. New England Journal of Medicine 377, 1639–1647 (2017).

11. Kaiboriboon, K. et al. Epilepsy surgery in the United States: Analysis of data from the National Association of Epilepsy Centers. Epilepsy Res 116, 105–109 (2015).

12. Cross, J. H., Reilly, C., Delicado, E. G., Smith, M. L. & Malmgren, K. Epilepsy surgery for children and adolescents: evidence-based but underused. The Lancet Child & Adolescent Health 6, 484–494 (2022).

13. Punia, V. The State of Specialized Epilepsy Care in the States: Increased Access, New Tools, but the Same Dismal Underutilization of Epilepsy Surgery. Epilepsy Curr 22, 228–230 (2022).

14. Beatty, C. W., Lockrow, J. P., Gedela, S., Gehred, A. & Ostendorf, A. P. The Missed Value of Underutilizing Pediatric Epilepsy Surgery: A Systematic Review. Seminars in Pediatric Neurology 39, 100917 (2021).

15. Maroufi, S. F. et al. Laser interstitial thermal therapy versus open resective surgery for nontumoral epilepsy: systematic review and meta-analysis of comparative studies. Journal of Neurosurgery 1, 1– 12 (2026).

16. Pichardo-Rojas, D. et al. A comparative assessment of laser interstitial thermal therapy and open resective surgery for drug-resistant epilepsy: a meta-analysis of 3873 patients. Journal of Neurosurgery 144, 35–54 (2025).

17. Yossofzai, O. et al. Seizure outcome of pediatric magnetic resonance-guided laser interstitial thermal therapy versus open surgery: A matched noninferiority cohort study. Epilepsia 64, 114–126 (2023).

18. Harris, W. B. et al. Long-term outcomes of pediatric epilepsy surgery: Individual participant data and study level meta-analyses. Seizure: European Journal of Epilepsy 101, 227–236 (2022).

19. Li, H., Ji, S., Dong, B. & Chen, L. Seizure control after epilepsy surgery in early childhood: A systematic review and meta-analysis. Epilepsy & Behavior 125, 108369 (2021).

20. Widjaja, E. et al. Seizure outcome of pediatric epilepsy surgery. Neurology 94, 311–321 (2020).

21. Hect, J. L. et al. Stereotactic laser interstitial thermal therapy for the treatment of pediatric drug-resistant epilepsy: indications, techniques, and safety. Childs Nerv Syst 38, 961–970 (2022).

22. Zeller, S. et al. Current applications and safety profile of laser interstitial thermal therapy in the pediatric population: a systematic review of the literature. Journal of Neurosurgery: Pediatrics 28, 360–367 (2021).

23. Woods, S. B. et al. Magnetic resonance–guided laser interstitial thermal therapy for pediatric drug-resistant epilepsy: a pooled analysis and systematic review of the literature. Journal of Neurosurgery: Pediatrics 36, 702–713 (2025).

24. Winslow, N., Himstead, A. & Vadera, S. Revision LITT for Epilepsy: How likely are patients to get a second treatment if the first fails? Journal of Clinical Neuroscience 136, 111235 (2025).

25. De Witt, M. E., Almaguer-Ascencio, M., Petropoulou, K. & Tovar-Spinoza, Z. The use of stereotactic MRI-guided laser interstitial thermal therapy for the treatment of pediatric cavernous malformations: the SUNY Upstate Golisano Children’s Hospital experience. Childs Nerv Syst 39, 417–424 (2023).

26. Sheikh, S. R. et al. Cost-effectiveness of surgery for drug-resistant temporal lobe epilepsy in the US. Neurology 95, e1404–e1416 (2020).

27. Discount Rates for Cost-Effectiveness Analysis of Federal Programs. Federal Register https://www.federalregister.gov/documents/2026/03/11/2026-04726/discount-rates-for-cost-effectiveness-analysis-of-federal-programs (2026).

28. Wieser, H. G. et al. ILAE Commission Report. Proposal for a new classification of outcome with respect to epileptic seizures following epilepsy surgery. Epilepsia 42, 282–286 (2001).

29. Lu, V. M., Brown, E. C., Ragheb, J. & Wang, S. Repeat surgery for pediatric epilepsy: a systematic review and meta-analysis of resection and disconnection approaches. J Neurosurg Pediatr 30, 616– 623 (2022).

30. Arias, E., Xu, J., Betzaida, T.-V. & Bastian, B. U.S. State Life Tables, 2021. https://stacks.cdc.gov(2024) doi:10.15620/cdc/157499.

31. Choi, H. et al. Epilepsy Surgery for Pharmacoresistant Temporal Lobe Epilepsy: A Decision Analysis. JAMA 300, 2497–2505 (2008).

32. Widjaja, E., Papastavros, T., Sander, B., Snead, C. & Pechlivanoglou, P. Early economic evaluation of MRI-guided laser interstitial thermal therapy (MRgLITT) and epilepsy surgery for mesial temporal lobe epilepsy. PLOS ONE 14, e0224571 (2019).

33. Salanova, V., Markand, O. & Worth, R. Temporal lobe epilepsy surgery: outcome, complications, and late mortality rate in 215 patients. Epilepsia 43, 170–174 (2002).

34. Sperling, M. R., Feldman, H., Kinman, J., Liporace, J. D. & O’Connor, M. J. Seizure control and mortality in epilepsy. Annals of Neurology 46, 45–50 (1999).

35. Annegers, J. F. et al. Epilepsy, vagal nerve stimulation by the NCP system, mortality, and sudden, unexpected, unexplained death. Epilepsia 39, 206–212 (1998).

36. Nilsson, L., Ahlbom, A., Farahmand, B. Y. & Tomson, T. Mortality in a population-based cohort of epilepsy surgery patients. Epilepsia 44, 575–581 (2003).

37. Guinle, M. I. B. et al. Approach, complications, and outcomes for 37 consecutive pediatric patients undergoing laser ablation for medically refractory epilepsy at Stanford Children’s Health. Journal of Neurosurgery: Pediatrics 33, 1–11 (2023).

38. Yossofzai, O. et al. Seizure outcome of pediatric magnetic resonance-guided laser interstitial thermal therapy versus open surgery: A matched noninferiority cohort study. Epilepsia 64, 114–126 (2023).

39. Drane, D. L. MRI-Guided stereotactic laser ablation for epilepsy surgery: Promising preliminary results for cognitive outcome. Epilepsy Research 142, 170–175 (2018).

40. Drane, D. L. et al. Superior Verbal Memory Outcome After Stereotactic Laser Amygdalohippocampotomy. Front Neurol 12, 779495 (2021).

