## Supplemental Table 1 for "Epilepsy Surgery vs Medical Management for Pediatric Drug-Resistant Focal Epilepsy"

**Supplemental Table 1. Sources for Model Input Parameters**

| **Parameter** | **Source(s)** |
| --- | --- |
| ***General Parameters*** |  |
| Annual discount rate | 1 |
| Starting cohort age | 2 |
| ***Initial Surgery Clinical Probabilities*** |  |
| Perioperative mortality - MRgLITT and open resection | Assumption |
| **MRgLITT** |  |
| Surgical complication | 2–7 |
| Permanent neurologic deficit | 2,4,5,7,8 |
| Seizure outcomes | 2–4,6,8,9 |
| Revision surgery | 2,6,8–10 |
| **Open Resection** |  |
| Surgical complication | 2–4,7,11 |
| Permanent neurologic deficit | 2,4,7 |
| Seizure outcomes | 2–4,12–14 |
| Revision surgery | 2,10,15,16 |
| **Revision Surgery** |  |
| Surgical complication | 17–21 |
| Seizure outcome (freedom) | 2,15–17,22 |
| **Best Medical Management** |  |
| Seizure outcome (freedom) | Calculated |
| ***Annual Seizure State Transition Probabilities*** |  |
| All seizure recurrence and remission probabilities | 23 |
| ***Health State Utilities*** |  |
| All utility values for all health states | 23 |
| ***Standardized Mortality Ratios (SMR)*** |  |
| All SMRs | 24–27 |
| ***Direct Medical Costs*** |  |
| All costs^A^ | 28 |
| ***Etiology-Specific Probability of Seizure Freedom*** |  |
| FCD – MRgLITT | 9,13,29–31 |
| FCD – Open resection | 13,14,29,30 |
| Tumor - MRgLITT | 2,9 |
| Tumor – Open resection | 2,9,14,32,33 |
| MTS - MRgLITT | 2,34,35 |
| MTS – Open resection | 14,35 |
| Cavernoma - MRgLITT | 9,36–38 |
| Cavernoma – Open resection | 37,39,40 |
| Abbreviations: MRgLITT = MRI-guided laser interstitial thermal therapy; FCD = focal cortical dysplasia; MTS = mesial temporal sclerosis | |
| ^A^All costs inflated and converted to 2025 US dollars (USD) | |

**References**

1.	Discount Rates for Cost-Effectiveness Analysis of Federal Programs. Federal Register. March 11, 2026. Accessed July 8, 2026. https://www.federalregister.gov/documents/2026/03/11/2026-04726/discount-rates-for-cost-effectiveness-analysis-of-federal-programs

2.	Yossofzai O, Stone S, Madsen J, et al. Seizure outcome of pediatric magnetic resonance-guided laser interstitial thermal therapy versus open surgery: A matched noninferiority cohort study. *Epilepsia*. 2023;64(1):114-126. doi:10.1111/epi.17451

3.	Maroufi SF, Fallahi MS, Amirkhani N, et al. Laser interstitial thermal therapy versus open resective surgery for nontumoral epilepsy: systematic review and meta-analysis of comparative studies. *Journal of Neurosurgery*. 2026;1(aop):1-12. doi:10.3171/2025.8.JNS25496

4.	Pichardo-Rojas D, Espinosa-Cantú CB, Valenzuela-Rangel AF, et al. A comparative assessment of laser interstitial thermal therapy and open resective surgery for drug-resistant epilepsy: a meta-analysis of 3873 patients. *Journal of Neurosurgery*. 2025;144(1):35-54. doi:10.3171/2025.4.JNS25386

5.	Zeller S, Kaye J, Jumah F, et al. Current applications and safety profile of laser interstitial thermal therapy in the pediatric population: a systematic review of the literature. *Journal of Neurosurgery: Pediatrics*. 2021;28(3):360-367. doi:10.3171/2021.2.PEDS20721

6.	Woods SB, Shields LBE, Kuruvilla A, et al. Magnetic resonance–guided laser interstitial thermal therapy for pediatric drug-resistant epilepsy: a pooled analysis and systematic review of the literature. *Journal of Neurosurgery: Pediatrics*. 2025;36(6):702-713. doi:10.3171/2025.5.PEDS25121

7.	Slingerland AL, Chua MMJ, Bolton J, et al. Stereoelectroencephalography followed by combined electrode removal and MRI-guided laser interstitial thermal therapy or open resection: a single-center series in pediatric patients with medically refractory epilepsy. *Journal of Neurosurgery: Pediatrics*. 2022;31(3):206-211. doi:10.3171/2022.11.PEDS22262

8.	Wittayanakorn N, Wong GM, Teti SA, et al. Outcomes of reoperation following failed laser ablation surgery for epilepsy in pediatric patients. *Journal of Neurosurgery: Pediatrics*. 2025;35(4):369-378. doi:10.3171/2024.11.PEDS24410

9.	Winslow N, Himstead A, Vadera S. Revision LITT for Epilepsy: How likely are patients to get a second treatment if the first fails? *Journal of Clinical Neuroscience*. 2025;136:111235. doi:10.1016/j.jocn.2025.111235

10.	Hsieh JK, Sundar SJ, Lu E, et al. Determinants of epileptogenic zone identification and seizure outcome in children with refractory epilepsy undergoing stereoelectroencephalography. *Journal of Neurosurgery: Pediatrics*. 2023;32(5):535-544. doi:10.3171/2023.5.PEDS22548

11.	Kumar RM, Koh S, Knupp K, Handler MH, O’Neill BR. Surgery for infants with catastrophic epilepsy: an analysis of complications and efficacy. *Childs Nerv Syst*. 2015;31(9):1479-1491. doi:10.1007/s00381-015-2759-6

12.	Harris WB, Brunette-Clement T, Wang A, et al. Long-term outcomes of pediatric epilepsy surgery: Individual participant data and study level meta-analyses. *Seizure: European Journal of Epilepsy*. 2022;101:227-236. doi:10.1016/j.seizure.2022.08.010

13.	Li H, Ji S, Dong B, Chen L. Seizure control after epilepsy surgery in early childhood: A systematic review and meta-analysis. *Epilepsy & Behavior*. 2021;125:108369. doi:10.1016/j.yebeh.2021.108369

14.	Widjaja E, Jain P, Demoe L, Guttmann A, Tomlinson G, Sander B. Seizure outcome of pediatric epilepsy surgery. *Neurology*. 2020;94(7):311-321. doi:10.1212/WNL.0000000000008966

15.	Cohen NT, Ziobro JM, Depositario-Cabacar DF, et al. Measure thrice, cut twice: On the benefit of reoperation for failed pediatric epilepsy surgery. *Epilepsy Research*. 2020;161:106289. doi:10.1016/j.eplepsyres.2020.106289

16.	Muthaffar O, Puka K, Rubinger L, et al. Reoperation after failed resective epilepsy surgery in children. *Journal of Neurosurgery: Pediatrics*. 2017;20(2):134-140. doi:10.3171/2017.3.PEDS16722

17.	Lu VM, Brown EC, Ragheb J, Wang S. Repeat surgery for pediatric epilepsy: a systematic review and meta-analysis of resection and disconnection approaches. *J Neurosurg Pediatr*. 2022;30(6):616-623. doi:10.3171/2022.9.PEDS22344

18.	Bower RS, Wirrell EC, Eckel LJ, Wong-Kisiel LC, Nickels KC, Wetjen NM. Repeat resective surgery in complex pediatric refractory epilepsy: lessons learned. *Journal of Neurosurgery: Pediatrics*. 2015;16(1):94-100. doi:10.3171/2014.12.PEDS14150

19.	Kunz M, Karschnia P, Borggraefe I, Noachtar S, Tonn JC, Vollmar C. Seizure-free outcome and safety of repeated epilepsy surgery for persistent or recurrent seizures. *Journal of Neurosurgery*. 2022;138(1):9-18. doi:10.3171/2022.4.JNS212753

20.	Ramantani G, Strobl K, Stathi A, et al. Reoperation for refractory epilepsy in childhood: a second chance for selected patients. *Neurosurgery*. 2013;73(4):695-704. doi:10.1227/NEU.0000000000000081

21.	Surges R, Elger CE. Reoperation after failed resective epilepsy surgery. *Seizure*. 2013;22(7):493-501. doi:10.1016/j.seizure.2013.04.020

22.	Krucoff MO, Chan AY, Harward SC, et al. Rates and predictors of success and failure in repeat epilepsy surgery: A meta-analysis and systematic review. *Epilepsia*. 2017;58(12):2133-2142. doi:10.1111/epi.13920

23.	Choi H, Sell RL, Lenert L, et al. Epilepsy Surgery for Pharmacoresistant Temporal Lobe Epilepsy: A Decision Analysis. *JAMA*. 2008;300(21):2497-2505. doi:10.1001/jama.2008.771

24.	Salanova V, Markand O, Worth R. Temporal lobe epilepsy surgery: outcome, complications, and late mortality rate in 215 patients. *Epilepsia*. 2002;43(2):170-174. doi:10.1046/j.1528-1157.2002.33800.x

25.	Sperling MR, Feldman H, Kinman J, Liporace JD, O’Connor MJ. Seizure control and mortality in epilepsy. *Annals of Neurology*. 1999;46(1):45-50. doi:10.1002/1531-8249(199907)46:1<45::AID-ANA8>3.0.CO;2-I

26.	Annegers JF, Coan SP, Hauser WA, Leestma J, Duffell W, Tarver B. Epilepsy, vagal nerve stimulation by the NCP system, mortality, and sudden, unexpected, unexplained death. *Epilepsia*. 1998;39(2):206-212. doi:10.1111/j.1528-1157.1998.tb01360.x

27.	Nilsson L, Ahlbom A, Farahmand BY, Tomson T. Mortality in a population-based cohort of epilepsy surgery patients. *Epilepsia*. 2003;44(4):575-581. doi:10.1046/j.1528-1157.2003.03302.x

28.	Widjaja E, Papastavros T, Sander B, Snead C, Pechlivanoglou P. Early economic evaluation of MRI-guided laser interstitial thermal therapy (MRgLITT) and epilepsy surgery for mesial temporal lobe epilepsy. *PLOS ONE*. 2019;14(11):e0224571. doi:10.1371/journal.pone.0224571

29.	Hale AT, Sen S, Haider AS, et al. Open Resection versus Laser Interstitial Thermal Therapy for the Treatment of Pediatric Insular Epilepsy. *Neurosurgery*. 2019;85(4):E730. doi:10.1093/neuros/nyz094

30.	Jain P, Ochi A, McInnis C, et al. Surgical outcomes in children with bottom-of-sulcus dysplasia and drug-resistant epilepsy: a retrospective cohort study. *Journal of Neurosurgery: Pediatrics*. 2021;28(3). doi:10.3171/2021.2.PEDS20967

31.	Yang B, Zhang C, Wang X, et al. Laser interstitial thermal therapy in the management of bottom-of-sulcus dysplasia-related epilepsy. *Annals of Clinical and Translational Neurology*. 2025;12(1):110-120. doi:10.1002/acn3.52258

32.	Englot DJ, Berger MS, Barbaro NM, Chang EF. Factors associated with seizure freedom in the surgical resection of glioneuronal tumors. *Epilepsia*. 2012;53(1):51-57. doi:10.1111/j.1528-1167.2011.03269.x

33.	Katlowitz KA, Athukuri P, Sharma H, et al. Seizure outcomes after resection of primary brain tumors in pediatric patients: a systematic review and meta-analysis. *J Neurooncol*. 2023;164(3):525-533. doi:10.1007/s11060-023-04446-9

34.	Brotis AG, Giannis T, Paschalis T, Kapsalaki E, Dardiotis E, Fountas KN. A meta-analysis on potential modifiers of LITT efficacy for mesial temporal lobe epilepsy: Seizure-freedom seems to fade with time. *Clinical Neurology and Neurosurgery*. 2021;205:106644. doi:10.1016/j.clineuro.2021.106644

35.	Kohlhase K, Zöllner JP, Tandon N, Strzelczyk A, Rosenow F. Comparison of minimally invasive and traditional surgical approaches for refractory mesial temporal lobe epilepsy: A systematic review and meta-analysis of outcomes. *Epilepsia*. 2021;62(4):831-845. doi:10.1111/epi.16846

36.	De Witt ME, Almaguer-Ascencio M, Petropoulou K, Tovar-Spinoza Z. The use of stereotactic MRI-guided laser interstitial thermal therapy for the treatment of pediatric cavernous malformations: the SUNY Upstate Golisano Children’s Hospital experience. *Childs Nerv Syst*. 2023;39(2):417-424. doi:10.1007/s00381-022-05701-6

37.	Bosisio L, Cognolato E, Nobile G, et al. Surgical treatment of cavernous malformation-related epilepsy in children: case series, systematic review, and meta-analysis. *Neurosurg Rev*. 2024;47(1):251. doi:10.1007/s10143-024-02491-0

38.	Mirone G, Meccariello G, Tengattini F, et al. Magnetic resonance-guided laser interstitial thermal therapy (MRgLITT) for paediatric intracranial cavernous malformations: case series and review of the literature. *Childs Nerv Syst*. 2025;41(1):183. doi:10.1007/s00381-025-06839-9

39.	Gao X, Yue K, Sun J, et al. A systematic review and meta-analysis of surgeries performed for cerebral cavernous malformation-related epilepsy in pediatric patients. *Front Pediatr*. 2022;10. doi:10.3389/fped.2022.892456

40.	Moran NF, Fish DR, Kitchen N, Shorvon S, Kendall BE, Stevens JM. Supratentorial cavernous haemangiomas and epilepsy: a review of the literature and case series. *Journal of Neurology, Neurosurgery & Psychiatry*. 1999;66(5):561-568. doi:10.1136/jnnp.66.5.561
